# Computed Tomography Findings and Short-term follow-up with Novel Coronavirus Pneumonia

**DOI:** 10.1101/2020.04.02.20042614

**Authors:** Shi Qi, Hui Guo, Hua Shao, Siqin Lan, Yuanlin He, Maijudan Tiheiran, Hongjun Li

## Abstract

**Objective:** To assess the characteristics of computed tomography (CT) features and changes in CT monitoring in patients with novel coronavirus pneumonia (NCP).

**Methods:** In this retrospective, two-center study, we reviewed the medical records of 57 patients with NCP in CT from January 21 to February 12, 2020. Cases were confirmed by the results of nucleic acid test positive, and were analyzed for demographic, clinical, and CT features.

**Results:** Of the 57 patients, 31cases were male, and 45.6% were female. The average age was 46.5 ± 15.8 years. Patients had fever (84.2%), cough (49.1%), weak (31.6%), muscle ache (17.5%), shortness of breath (12.3%). The distribution of abnormality was a subpleural lesions in 51 cases, with 96.5% ground-glass opacity (GGO) and 68.4% consolidation. Another observation reveals 45.6% fibrosis, 33.3% lymph node enlargement, 21.1% pleural thickening, 17.5% small nodule, 7.0% white lung, 5.3% emphysema, and 3.5% bronchiectasis. Importantly, the group of men had more septal thickening and air trapping than the female group (p<0.05); Compared with the younger, the elderly had higher of subpleural lesion, interlobular septal thickening and pleural thickening (p<0.05). In the first monitoring, there were 37.3% improvement, 60.8% progress. In the second monitoring, there were 55% improvement, 35% progress. The improvement rate during the third follow-up visit was 100%.

**Conclusions:** CT features and CT dynamic observation play a vital role in the diagnosis and treatment with NCP. It is conducive to early diagnosis, deepen the knowledge of NCP and accumulate experience.

**Key points:** Bilateral and subpleural regions on CT are common in patients with novel coronavirus pneumonia (NCP), which were more extensive GGOs than consolidation.

At CT, NCP shows GGOs without or with consolidation, which most of the patients appear interlobular interstitium thickening, vascular bundle thickening, air bronchogram, and fibrosis. In patients with fever in close contact with eruption areas of 2019-nCoV, familiarity with the CT findings may help with early diagnosis, early isolation, and management.

## Introduction

In December 2019, acute respiratory disease broke out in Wuhan City, Hubei Province, China, now known as novel coronavirus pneumonia (NCP)^1-7^. The disease has been rapidly spread to other districts and other countries by Wuhan. As of February 16, 2020, a total of 26 states and five continents have reported cases^6^. 2019-nCoV not only is it an acute disease, but it is also an effective communication between people through coughing, sneezing or direct contact^8^. Although most of the time patients have no or slight clinical manifestations of infection with 2019-nCoV, the imaging performance is usually good and serious. Lung injury caused by 2019-nCoV is one of the leading clinical signs and affects the prognosis directly. Imaging CT plays an essential role in the diagnosis and evaluation of patients with NCP. Although the national CT strategy is documented, those studies focus mainly on the epidemiology, clinical treatment, and disease treatment^7, 9-11^. So far, we have identified some of the studies concerning CT results ^12-16^. There’s very little research on the CT quality and the NCP CT scan results. The current retrospective study was intended to evaluate the thin-slice CT features and CT-follow-up results in NCP patients.

## Material and Methods

### Patients

In this retrospective binocular center study, 57 patients with NCP were examined on plain CT from January 21 to Febuary 15, 2020 in the First Hospital of Xinjiang Medical University and the Affiliated Beijing Youan Hospital of Beijing Capital Medical University. The cases were confirmed with a positive nucleic acid test and analyzed for demographic, clinical and CT features. Follow up to February 19, 2020. Ethical approval for the study was obtained from the Ethical Review Committee of the First Affiliated Hospita of Xinjiang Medical University (No.2020120-06). The present study abandoned the need for individual consent for retrospective observation studies.

### CT scans

CT scans were created using a 64-speed multi-scan system (High Speed; GE Medical Systems, Milwaukee, WI, USA, and Light Speed; GE Medical Systems, Milwaukee, WI, USA). Nonenhanced CT had been processed with numbers: 120 kVp, 150 mA, 5-mm thick. Thin-section CT images have been reconstructed using 1-mm or 1.25-mm collimation, which have been evaluated using a lung window, 1600-window width and 600-window level, and then send to the workstation (ADW 4.2; GE Medical Systems, Milwaukee, WI, USA) by analysis.

### Image interpretation

All CT images were independently evaluated by two radiologists (GH and QS with 15 and 20 years’ CT experience, respectively). Both radiologists had to agree on the anomaly.

Taking into account standard morphological descriptors based on the recommendations of the Fleischner Society Nomenclature Committee^17^ and other study^18^, the examination of thin-slice CT abnormalities (unilateral lung, unilateral lung or pulmonary segment) included distribution of lesions, ground-glass opacity (GGO), number (single or two or multiple), GGO with consolidation, consolidation, air trapping, air bronchogram, vascular bundle thickening, fibrosis, interlobular septal thickening, lymph node enlargement (≥5 mm), pleural thickening, crazy-paving, small nodule, halo sign, white lung, bronchiectasis, emphysema. The assessment of the extent of lung involvement was based on segments of pulmonary anatomy: 10 segments in the right lung and ten segments in the left lung (2 segments were considered in the apicoposterius segment left upper lobe, and two segments were considered in the inferior front segment of the left lower lobe).

Follow-up of NCP patients was improved, partial improvement - progress, and progression. The improvement was defined as absorption or/and area reduction of one or/and two or multiple GGOs or/and consolidation. Partial improvement - progression was defined as local absorption or/and partial reduction of GGOs or/and consolidation. Progression was defined as area increasing or/and lesion as addition of GGOs or/and consolidation, and white lung may even occur.

### Statistical analysis

We collected date on age, symptoms, CT finding. All data are expressed as the percentage. We collected the data on age and clinical time. All data are expressed as means ± standard deviation (SD). The Chi-square analysed some of the differences between the two groups. A P < 0.05 was considered a statistically significant difference.

## Results

### Baseline characteristics of patients

The basic characteristics of NCP patients are shown in Table 1. Of the 57 patients, 31cases were male, and 45.6% were female. The average age of the patients was 46.5 ± 15.8 years (age range 1-78 years). 2 patients were children (aged one years and 16 years). The time from the onset of symptoms to hospital presentation ranged from 2 to 10 days (mean time 5.0 ± 1.9 days). The most common symptom was fever (84.2%), followed by cough (49.1%), weak (31.6%), muscle soreness (17.5%), shortness of breath (12.3%), respectively. The temperature of no less than 38° had 32 cases.

**Table 1.**
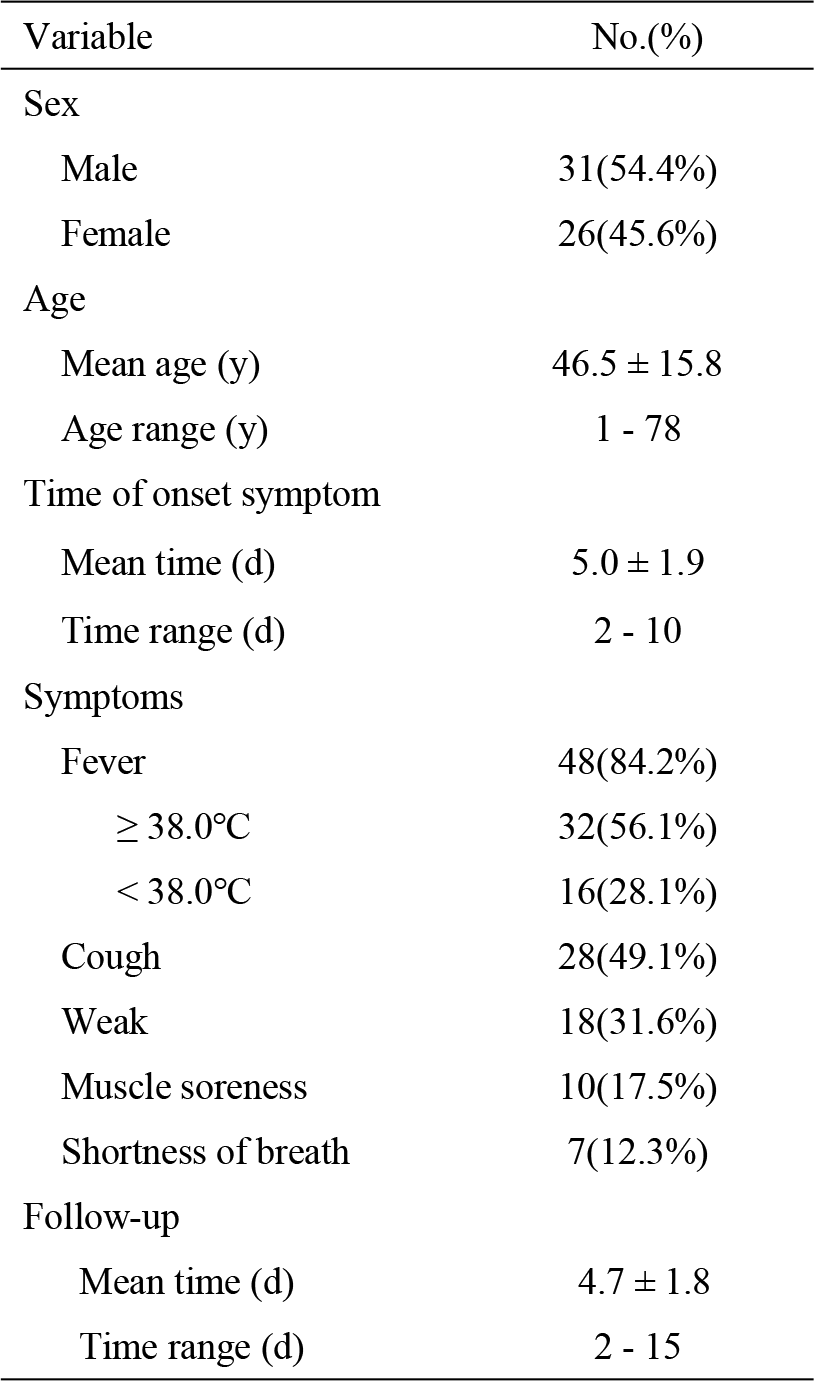
Demographic and clinical characteristics of patients with novel coronavirus pneumonia

| Variable | No.(%) |
| --- | --- |
| Sex |  |
| Male | 31(54.4%) |
| Female | 26(45.6%) |
| Age |  |
| Mean age (y) | 46.5 ± 15.8 |
| Age range (y) | 1 - 78 |
| Time of onset symptom |  |
| Mean time (d) | 5.0 ± 1.9 |
| Time range (d) | 2 - 10 |
| Symptoms |  |
| Fever | 48(84.2%) |
| ≥ 38.0°C | 32(56.1%) |
| < 38.0°C | 16(28.1%) |
| Cough | 28(49.1%) |
| Weak | 18(31.6%) |
| Muscle soreness | 10(17.5%) |
| Shortness of breath | 7(12.3%) |
| Follow-up |  |
| Mean time (d) | 4.7 ± 1.8 |
| Time range (d) | 2 - 15 |

### CT findings

All patients underwent CT on the day of admission. 51 CT scans were first repeated in the hospital, and 20 patients, one case was investigated in the second follow-up, third follow-up, as appropriate.

The distribution of NCP lesions on chest CT patterns is shown in Table 2. Of the 57 patients, 15 (25.3%) were unidirectional except for one without abnormality: right lung in 6 (10.5%) and left lung in 9 (15.8%). The lesions were bilateral in 41 (71.9%) patients. The distribution of anomalies was predominantly subpleural lesions in 51 (89.5%) patients (Fig. 1). The most lesions were located in the anteromedial basal (63.2%) from the lower lobe of the left lung, followed by the lateral basal (59.6%) of the lower lobe of the right lung, medial basal (47.4%) of the lower lobe of the left lung, etc. (summarized in Table 2). One lung had one segment and two segments involved in 7 cases. Parenchymal changes primarily represented GGOs in the lung. The three categories of GGO are single (8.8%), two (15.8%), multiple (48.2%), respectively.

**Table 2.** Distribution of the lesions in patients with NCP

| CT finding | Right lung<br>(No., %) | Left lung<br>(No., %) |
| --- | --- | --- |
| Superior lobe |  |  |
| Apical | 19(33.3%) | - |
| Posterior (Apicoposterior) | 24(42.1%) | 22(38.6%) |
| Anterior | 13(22.8%) | 12(21.1%) |
| Middle lobe (Superior lobe ) |  |  |
| Lateral (Superior lingular) | 17(29.8%) | 18(31.6%) |
| Medial (Inferior lingular) | 17(29.8%) | 17(29.8%) |
| Inferior lobe |  |  |
| Superior | 26(45.6%) | 25(43.9%) |
| Anterior basal | 11(19.3%) | 17(29.8%) |
| Medial basal | 24(42.1%) | 27(47.4%) |
| Lateral basal (Anteromedial basal) | 34(59.6%) | 36(63.2%) |
| Posterior basal | 26(45.6%) | 22(38.6%) |
| Unilateral lung | 6(10.5%) | 9(15.8%) |
| Subpleural lesion | 24(42.1%) | 27(47.4%) |
| Single GGO | 2(3.5%) | 3(5.3%) |
| Two GGO | 4(7.0%) | 5(8.8%) |
| Multiple GGO | 41(71.9%) | 39(68.4%) |

Notes: NCP = Novel coronavirus pneumonia; GGO = Ground-glass opacity

**Figure 1.**
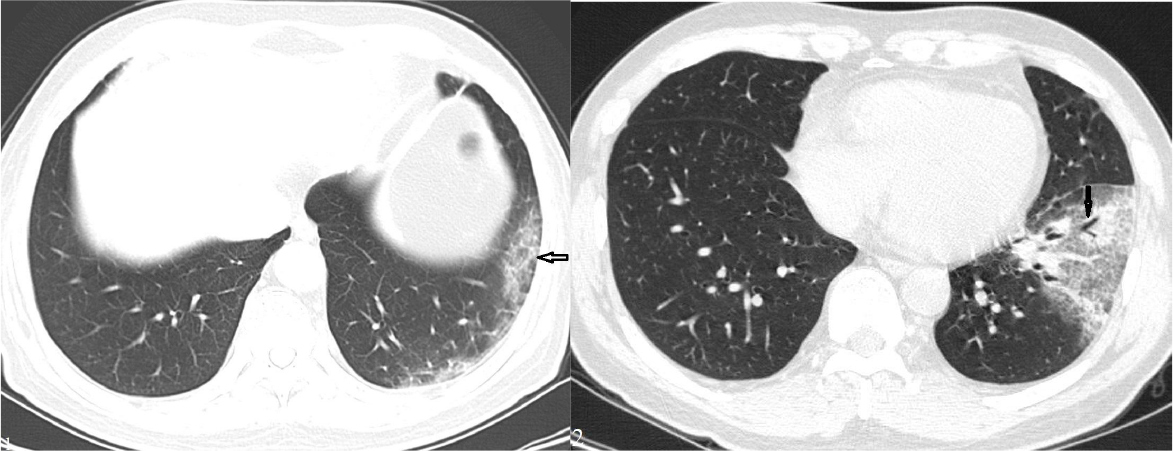
Male, 72 years old. The CT scan showed subpleural GGO in the lower lobe of the left lung (arrow).

Results of the CT scan in patients with NCP are shown in Table 3. The most common thin - slice CT finding were GGOs of the lung (Fig. 1), detecting in 55 of the 57 (96.5%) cases, followed by fibrosis (45.6%), consolidation (36.8%), lymph node enlargement (33.3%), pleural thickening(21.1%), small nodule (17.5%), white lung (7.0%), emphysema (5.3%), bronchiectasis (3.5%) (Fig. 2). In some patients, GGO was associated with other lesions, including GGO with air bronchogram (57.9%) (Fig. 2), vascular bundle thickening (45.6%) (Fig. 3A), interlobular septal thickening (40.4%) (Fig. 4A), consolidation (31.6%) (Fig. 5), air trapping (22.8%), crazy-paving (19.3%) (Fig. 3B), and halo sign (14.0%) (Fig. 3C).

**Table 3.** CT imaging findings in patients with novel coronavirus

| CT finding | Examination hospitalized<br>(n=57) (No., %) |
| --- | --- |
| GGO | 55(96.5%) |
| GGO |  |
| Air bronchogram | 33(57.9%) |
| Vascular bundle thickening | 26(45.6%) |
| Interlobular septal thickening | 23(40.4%) |
| Consolidation | 18(31.6%) |
| Air trapping | 13(22.8%) |
| Crazy-paving sign | 11(19.3%) |
| Halo sign | 8(14.0%) |
| Fibrosis | 26(45.6%) |
| Consolidation | 21(36.8%) |
| Lymph node enlargement | 19(33.3%) |
| Pleural thickening | 12(21.1%) |
| Small nodule | 10(17.5%) |
| White lung | 4(7.0%) |
| Emphysema | 3(5.3%) |
| Bronchiectasis | 2(3.5%) |

Note: GGO=Ground-glass opacity

**Figure 2.** Male, 48 years old. The CT scan showed subpleural GGO with air bronchogram in the lower lobe of the left lung (arrow).

**Figure 3A-C.**
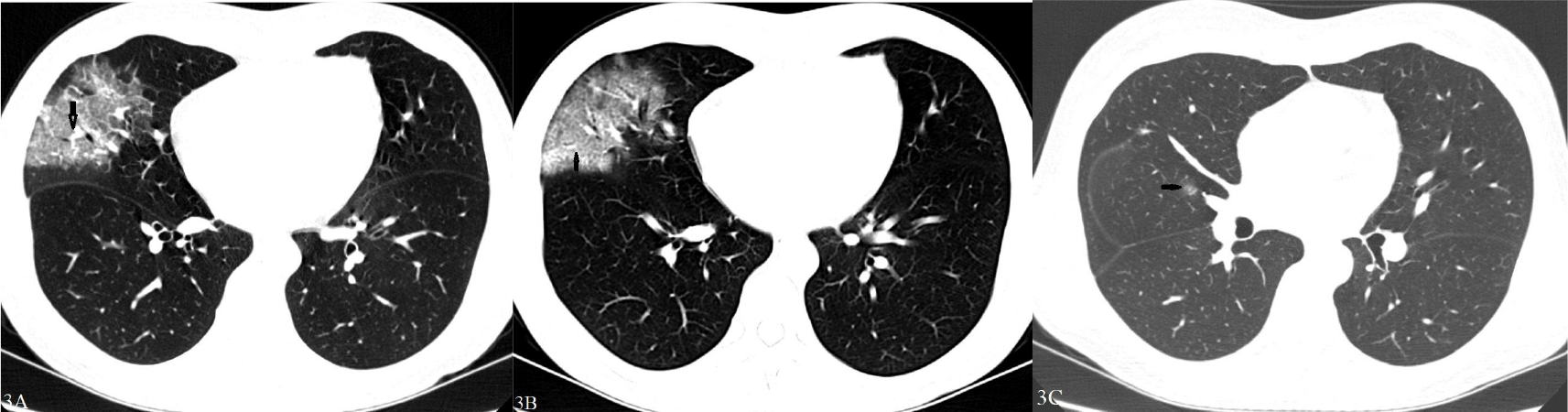
Female, 34 years old. (A) The CT scan showed subpleural GGO with vascular bundle thickening in the middle lobe of the right lung (arrow). (B) The CT scan showed subpleural GGO with crazy-paving in the middle lobe of the right lung (arrow). (C) The CT scan showed a small nodule with halo sign in the superior lobe of the right lung (arrow).

**Figure 4A-C.**
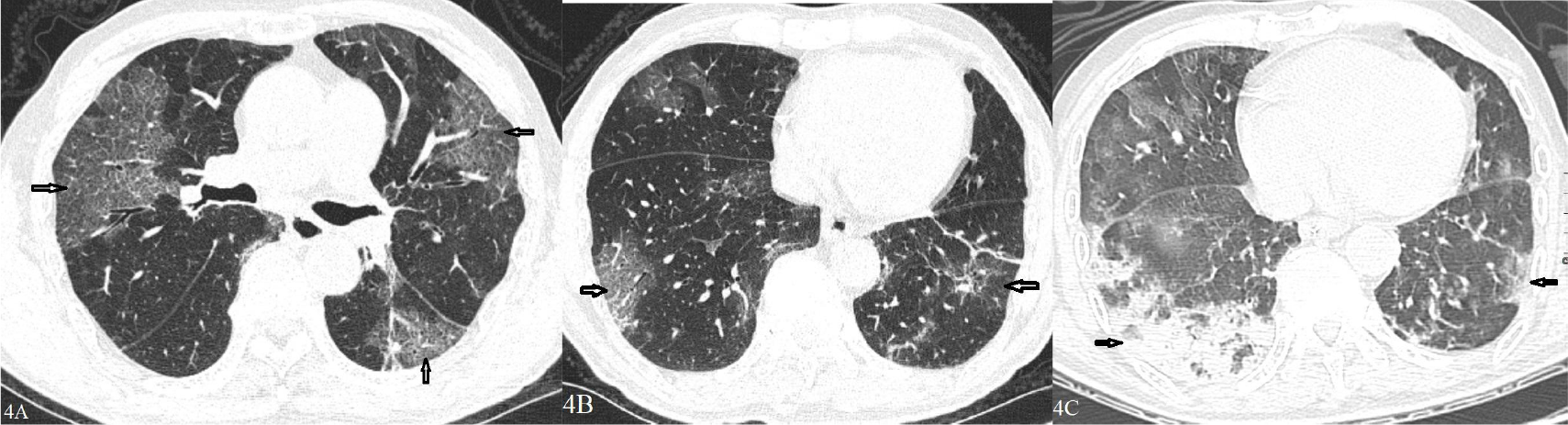
Male, 75 years old. (A) The CT scan showed subpleural GGO with interlobular septal thickening in the bilateral lung (arrow). (B) The CT scan showed multiple patchy areas of GGO in the bilateral lung (arrow). (C) Follow-up CT images on day 12 after admission showed an overlap of organizing pneumonia with diffuse alveolar damage in that it is more diffuse and associated with underlying reticulation. prominent progression with increased size and density of the lesions, and with more consolidations (arrow). Figure C also shows air bronchogram and fibrosis. Interlobular septal thickening does not seem to be a major component.

**Figure 5.**
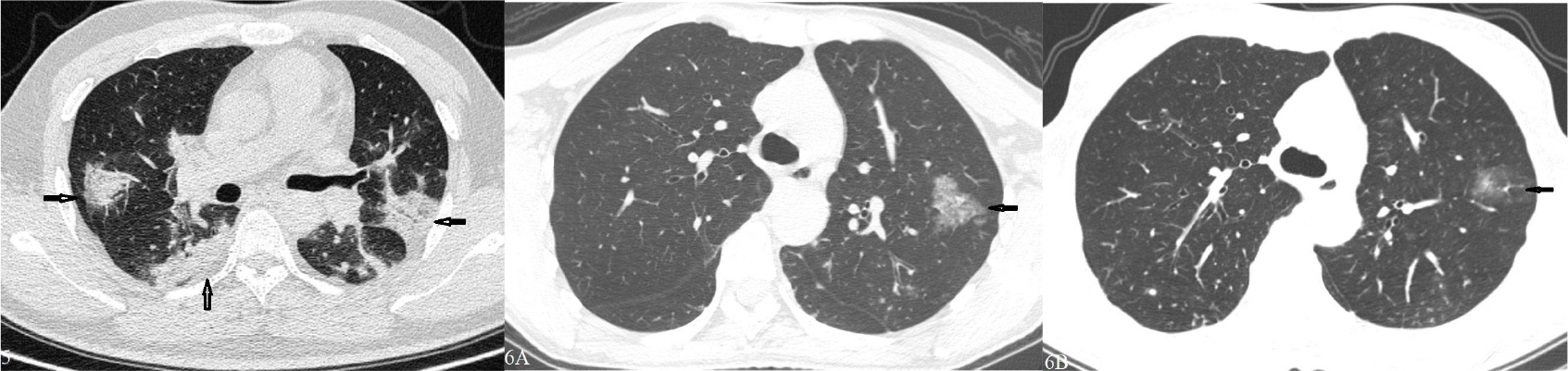
Male, 45 years old. The CT scan showed subpleural consolidation in the superior lobe of the bilateral lung (arrow). Figure 6A-B. Female, 45 years old. (A) The CT scan showed single GGO in the superior lobe of the left lung (arrow). (B) Follow-up CT images on day 4 after admission showed prominent improvement with decreased size and density of the lesions in the superior lobe of the left lung (arrow).

### Comparing CT characteristics between the two groups

We divided these patients into a group of man and female (Table 4). There were more interlobular septal thickening in the male group (54.8%) than in the female group (23.1%) (p=0.015). There were significantly more air trapping in the male group (35.5%) than in the female group (7.7%) (p=0.013).

**Table 4.** Comparing CT finding between male group and female group

| CT finding | Male group (n=31)<br>(No., %) | Female group (n=26)<br>(No., %) | $\chi^2$-value | P-value |
| --- | --- | --- | --- | --- |
| Interlobular septall thickening | 17 (54.8%) | 6 (23.1%) | 5.927 | 0.015 |
| Air trapping | 11 (35.5%) | 2 (7.7%) | 6.204 | 0.013 |

We stratified the patients into two age groups (Table 5), as age ≤ 50 years and age > 50 years.

**Table 5.** Comparing CT finding between the elder group and the younger group.

| CT finding | Elder group (n=22)<br>(No., %) | Younger group (n=35)<br>(No., %) | $\chi^2$-value | P-value |
| --- | --- | --- | --- | --- |
| Subpleural lesion | 22 (100%) | 29 (82.9%) | 4.215 | 0.040 |
| Interlobular septal thickening | 13 (59.1%) | 10 (28.6%) | 5.228 | 0.022 |
| Lymph node enlargement | 8 (36.4%) | 4 (10.5%) | 5.054 | 0.025 |

We found more subpleural lesions (including GGO, consolidation, small nodule) in the elder group (100%) than in the younger group (82.9%) (p=0.040). In the elder group (59.1 %), there were significantly more interlobular septal thickening than in the younger group (28.6%). It was considerably more of pleural thickening in the elder group (36.4%) when compared to the younger group (11.4 %) (p=0.025).

### Follow-up changes by CT findings over time

Fifty-one patients were monitored, and the mean monitoring was 4.7 ± 1.8 days (range, 2 to 15 days). The mean time from hospital to the first monitoring was three days (range, 2 to 11 days). Monitoring of 1-time period was 51 patients, 20 cases of 2-times, 1 case of three-times.

The results of predominant CT findings showed the three types: improvement, partial improvement-progression, progression. When CT scanned the first follow-up in 51 patients, there were 37.3% improvement (Figure 6A-B), 2.0% partial improvement - progress, 60.8% progress (Figure 4B-C). The second CT follow-up performed twenty, and 11 cases, two patients and 7 cases showed an improvement, part improvement - progression, progression, respectively. Only one case was followed up on three CT scans, and 100% was improved.

## Discussion

This study shows that bilateral and subpleural regions on CT are common in patients with NCP, which were more extensive GGOs than consolidation. In one case, however, there was no abnormality, and the other had two small nodules in the lateral basal of the right lower lobe. Another observation is that more than one-third of patients reveal fibrosis, lymph node enlargement, and some patients may cause pleural thickening, small nodule, white lung, emphysema, and bronchiectasis. The most important thing is that the interlobular septal thickening and air trapping of the male group is more than that of the female group; In the elderly, the subpleural lesion, septal thickening, and pleural thickening increase much more than in the younger.

When faced with suspected cases of 2019-nCoV infection and need to identify NCP in time, we may encounter the following several reasons. First, patients with 2019-nCoV infection may develop asymptomatic or mild symptoms. Second, nucleic acid detections in patient’s respiratory samples may present negative results. Third, even in cases identified with 2019-nCoV, the result of nucleic acid testing may be given 24-hours later. In patients with fever in close contact with the eruption areas of 2019-nCoV, familiarity with the CT findings may assist in early diagnosis, early isolation, and management.

CT is useful for clarifying the abnormality of the patient’s lung and, especially, to describe the patterns and range of abnormality with coronavirus pneumonia^19-21^. In the current study, the predominance of GGOs and consolidation in the subpleural regions is remarkable. The GGOs and consolidation in such a distribution have been described as suggestive of a coronavirus^22,23^ and other pneumonia patterns^24,25^. However, they have different aspects. On CT images, centrilobular nodules, pseudocavitation, pneumatocele formation are commonly seen in H5N1 pneumonia^24^, but are not seen in NCP patients. During the disease, pleural effusions and cavitation can also be found in H5N1 pneumonia, but have not been developed in patients with NCP. In patients with severe rhinovirus pneumonia, bilateral patchy consolidation with multifocal GGO and septal thickening are noted^25^. Cavitation, pleural effusions, or lymphadenopathy are not characteristic manifestations of SARS pneumonia [26], but 33.3% increases in lymph node enlargements are observed in patients with NCP. A few patients may see interlobular septal thickening and pleural effusions in MERS pneumonia^22^, and pneumothorax and pleural effusion are more common in patients who have died than in those who have recovered^27^. In NCP patients, we haven’t seen pneumothorax, or pleural effusion, 40.4% interlobular septal thickening, 45.6% fibrosis, 45.6% vascular bundle thickening were often seen. In addition to the above findings, NCP also includes air trapping, crazy-paving, pleural thickening, halo sign, white lung, and emphysema. Another finding in this study is the presence of air bronchogram, air trapping, or bronchiectasis, all of which manifest as airway abnormalities. Air bronchogram, air trapping, bronchiectasis were found in 33 patients, 13 patients, two patients, respectively.

CT patterns of coronavirus pneumonia are associated with the pathogenesis of coronavirus infections. Most viral pneumonia of the imaging patterns share a similarity based on viridian, because viruses of the same miridae have a similar pathogenesis^28^. The pathological characteristics of NCP are probably interstitial interventions in alveolar walls and peribronchiolar, which are related to the pathogenesis of S-protein binding to ACE2 receptors in alveolar type III epithelial cells and subsequent damage. The physiological factors can show the damaged alveolar walls and septal, peribronchiolar, expansion congestion of alveolar-capillary, and distinctly organization infiltration in alveolar space^29^. At this stage, it is too early to draw reliable conclusions, but our opinion on 2019-nCoV can go through the same evolution as SARS-CoV.

We analyzed the CT characteristics between male and female groups. The interlobular septal thickening and air trapping in the male group were significantly higher that in female group. We also analyzed the CT characteristics between elder and younger groups. We discovered that there was significantly more subpleural lesions (including GGO, consolidation, and small nodule), interlobular septal thickening, and lymph node enlargement in the elder than in the younger group. These mechanisms are unclear and need further study.

At present, we analyzed in the first follow-up visit with an average duration of three days, improvement in 19 of the 51 cases (37.3%) and 60.8% progression. When we extended visits, 11 of the 20 patients (55%)performed better, 35.0% progression. It is interesting that two patients showed an increase in lymph nodes in the mediastinum during the second monitoring.

The present study is not without limitations. This study is a retrospective study of patients assessed by the two centers, including all selection questions and observation bias. Secondly, we need to further study the changes of a bigger sample in the CT features and CT follow-up with NCP.

In summary, CT characteristics and CT dynamic observation play a vital role in diagnosis and treatment with NCP, which is conducive to early diagnosis of the disease, our further understanding of the disease, and accumulation of experience.

## Data Availability

Statements
We state that all data are in the manuscript. Ethical approval for the study was obtained from the Ethical Review Committee of the First Affiliated Hospita of Xinjiang Medical University (No.2020120-06). The procedures were performed as per the guidelines of Declaration of Helsinki 2013.

## Abbreviation

NCP: Novel coronavirus pneumonia
CT: Computed tomography
GGO: Ground-glass opacity
SD: Standard deviation’
SARS: Severe acute respiratory syndrome MERS Middle East respiratory syndrome

